# A comprehensive benchmarking and validation study of cross-trait association methods

**DOI:** 10.1101/2022.09.07.22279671

**Authors:** Sander Lamballais, Gennady V. Roshchupkin, Raymond A. Poot, Steven A. Kushner, M. Arfan Ikram, Hieab H. H. Adams, Henning Tiemeier

**Affiliations:** Department of Clinical Genetics, Erasmus MC University Medical Center, Rotterdam, the Netherlands; Department of Epidemiology, Erasmus MC University Medical Center, Rotterdam, the Netherlands; Department of Radiology and Nuclear Medicine, Erasmus MC University Medical Center, Rotterdam, the Netherlands; Department of Cell Biology, Erasmus MC University Medical Center, Rotterdam, the Netherlands; Department of Psychiatry, Erasmus MC University Medical Center, Rotterdam,the Netherlands; Department of Psychiatry, Columbia University, New York, New York, USA; Department of Human Genetics, Radboud University Medical Center, Nijmegen, the Netherlands; Latin American Brain Health (BrainLat), Universidad Adolfo Ibáñez, Santiago, Chile; Department of Social and Behavioral Science, Harvard TH Chan School of Public Health, Boston, Massachusetts, USA

## Abstract

Cross-trait analyses are a powerful approach for refining our understanding of the genetic architectures underlying complex traits. Although many cross-trait association methods exist, a systematic evaluation of their performance is lacking. We compare true and false positive rates of several other methods using numerical (up to 300 traits) and genotype (up to 4 traits) simulations and introduce a new cross-trait association method, SumRank. For two traits, ConjFDR, SumRank, and GPA showed high true positive rates while maintaining false positive rates below 5%. SumRank and ASSET outperformed other methods for more than two traits. Most other methods had false positive rates well above 5%, with the false positive rates rising with the number of traits. Application of SumRank to eight psychiatric disorders yielded 658 novel loci, tripling the number of known loci. We discuss the differences in performance in cross-trait analyses as well as the risk of inflated false positives.

## Introduction

Pleiotropy occurs when a single gene or genetic locus influences more than one trait or disease. It explains how a limited number of genes can impact numerous traits simultaneously through shared biological pathways. By leveraging pleiotropy, insights into the genetic architecture of one disease can facilitate the elucidation of another. One powerful approach to detect pleiotropy is through cross-trait association analysis, which examines associations of genetic variants–such as single nucleotide polymorphism (SNPs)–with multiple traits. Such analyses capture both horizontal pleiotropy, where independent biological processes affect distinct traits, and vertical pleiotropy, in which one trait causally influences another. A recent study estimated that cross-trait associations span 57% of loci in the genome^1^, but this is likely an underestimation due to stringent statistical thresholds and our still-evolving understanding of pleiotropy. A complete understanding of pleiotropy thus requires the development and validation of robust cross-trait association methods capable of reliably prioritizing truly pleiotropic SNPs.

Genome-wide association studies (GWASs) are commonly used to uncover genetic architectures by evaluating SNP-trait associations^2^. A common approach to studying cross-trait associations with GWAS is by using multivariate GWAS, which analyzes individual-level data on multiple traits in the same study population. However, such data are often either unavailable or impractical to obtain. An alternative approach is to use summary-level methods, which employ only summary-level GWAS statistics and can be conducted on independent, non-overlapping populations. Summary-level methods estimate the probability that a SNP is pleiotropic by integrating effect sizes or p-values across different GWASs. Notable examples include *Fisher’s Combined Test* (FCT), *Cross-Phenotype Meta-Analysis* (CPMA)^3^, *ASsociation analysis based on subSETs* (ASSET)^4^, *Genetic analysis incorporating Pleiotropy and Annotation* (GPA)^5^, *conjunctional FDR* (conjFDR)^6^, *GWAS-pairwise* (GWAS-PW)^7^, and *PLeiotropic Analysis under COmposite null hypothesis* (PLACO)^8^ (**Table 1**). With the continuously growing number of GWASs available, these summary-level methods offer a straightforward and scalable approach to unravel the complex pleiotropic nature of the human genome.

**Table 1.** Overview of summary-level methods used to study cross-trait associations.

| Method | Year | Ref. | Traits | Hypotheses | Subset testing | Input | Output |
| --- | --- | --- | --- | --- | --- | --- | --- |
| FCT | 1925 | <sup>27</sup> | $\geq 2$ | $H_0$: The SNP is not associated with any trait | Possible | p-values | p-value given $H_0$ |
| CPMA | 2011 | <sup>3</sup> | $\geq 2$ | $H_0$: The SNP is not associated with any trait | Possible | p-values | p-value given $H_0$ |
| ASSET | 2012 | <sup>4</sup> | $\geq 2$ | $H_0$: The SNP is not associated with any subset of traits | Yes | $\beta$, $SE_\beta$, N | Subset of (positive/negative) associated traits and p-values |
| GPA | 2014 | <sup>5</sup> | 2 | 4-group model: $\pi_{00}$ (no association), $\pi_{10}$ (trait 1 only), $\pi_{01}$ (trait 2 only) and $\pi_{11}$ (pleiotropy) | No | p-values | Most likely model ($\pi_{00}$, $\pi_{10}$, $\pi_{01}$ or $\pi_{11}$) |
| ConjFDR | 2014 | <sup>6</sup> | 2 | $H_0$: The SNP is not associated with both traits | No | p-values | q-value for cross-trait association of SNP |
| GWAS-PW | 2016 | <sup>7</sup> | 2 | 4-group model: $\pi_{00}$ (no association), $\pi_{10}$ (trait 1 only), $\pi_{01}$ (trait 2 only) and $\pi_{11}$ (pleiotropy) | No / not relevant | Z, $r_g$ | Posterior probability for $\pi_{10}$, $\pi_{01}$ and $\pi_{11}$ |
| PLACO | 2020 | <sup>8</sup> | 2 | $H_0$: The SNP is not associated with both traits | No | p-values | q-value for cross-trait association of SNP |
| SumRank | 2023 | | $\geq 2$ | $H_0$: The SNP is not associated with all traits under study | Possible | p-values | p-value given $H_0$ |

Importantly, each of these methods relies on distinct statistical frameworks and assumptions, which lead to heterogenous performance across different scenarios. Moreover, most methods have only been tested under specific simulated conditions, and direct comparisons of their false positive and true positive rates remain lacking. Therefore, a comprehensive benchmark is needed to evaluate the performance of current approaches, thereby guiding the development of novel methods. Additionally, certain properties of one method could potentially be adapted to enhance others, paving the way for more powerful and reliable detection of cross-trait associations.

Here, we introduce SumRank as a novel method for cross-trait analysis and systematically compared its performance with existing summary-level methods in detecting valid cross-trait associations under diverse conditions. To benchmark the performance of the various methods, we created a comprehensive simulation framework. First, we tested the behavior of eight methods on simple, numerical simulations comparing two traits, focusing on false positive and true positive rates. Second, to approximate the complexity of real-world scenarios, we performed genotype simulations based on UK Biobank data, modeling genetically related traits. Third, given the growing interest in cross-trait associations across numerous traits, we extended the numerical simulations to 300 traits for a subset of methods, exploring approaches to properly account for false positives. These simulations were further corroborated by genotype simulations based on the UK Biobank data. Finally, we applied the cross-trait methods to GWAS summary data of eight psychiatric traits as a real-world application.

## Results

### Overview of methods for cross-trait associations

The approaches to detect pleiotropic SNPs greatly differ between the summary-level methods examined (**Table 1**), including in their hypotheses, the number of traits that can be analyzed simultaneously, their reliance on information from other SNPs, and the type of input data required. Still, the methods are unified in how they model the association between a SNP *i* and a trait Y*j* using a regression formula:

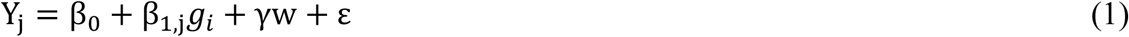

where β_1,j_ is the effect of SNP *i* on trait Y*j, g* is the genotype, γw is a non-genetic component, and ε is an error term. In this context, a pleiotropic SNP is defined as one where β_1,j_ ≠ 0 for multiple traits *j* among the *m* traits under consideration.

Here we will consider the differences between the cross-trait methods, starting first with the hypothesis. Methods such as FCT and CPMA test a non-composite null hypothesis where all β_1,1_ to β_1,m_ are zero. This means that any non-zero β_1,j_ will lead to a rejection of the null hypothesis, even if only one trait is associated. Alternatively, a composite null hypothesis posits that at least one β_1,j_ is zero, which leads to rejection of the null hypothesis if all traits are associated.

Second, although most cross-trait methods can only test associations across two traits, others can handle multiple traits simultaneously (**Table 1**). In the latter case, multiple null hypotheses can be formulated. The first is to test whether at least one of β_1,1_ … β_1,m_ are zero but only results in cross-trait associations when all traits are associated. The second approach, as is done with the ASSET method, is to test every possible subset of *m* traits and select the optimal subset, enabling the detection of cross-trait associations in specific trait combinations.

Third, most methods calculate the cross-trait associations for each SNP separately. However, methods like GWAS-PW instead incorporate information on linkage disequilibrium (LD) and optimize the model per defined genomic region, thus considering a broader genomic context.

Lastly, the methods differ in their input requirements – p-values, effect sizes, standard errors and genetic correlations (r_g_) – and their output– (un)adjusted p-values, posterior probabilities, or simply the selection of associated traits (**Table 1**).

In this study, we introduce SumRank, a novel combinatorial method for evaluating cross-trait associations based on p-values from genome-wide association study (GWAS) summary statistics. SumRank explicitly tests the composite hypothesis that a SNP is associated with all traits under study and can be extended to test for subsets of traits. We will compare various methods across a range of simulations and conditions, demonstrating how SumRank performs relative to existing methods.

### Numerical simulations for two traits

The differences between cross-trait methods are best illustrated by comparing their rejection plots for two traits (**Fig. 1a**). We present the plots for methods without (FCT and CPMA) and with a composite null hypothesis (SumRank and PLACO). When the SNP has low p-values in both GWASs (**Fig. 1a**, top-right in each plot), all methods correctly identify the SNP as pleiotropic. However, when the SNP is associated with only one trait (**Fig. 1a**, top-left or bottom-right in each plot), FCT and CPMA will still yield low p-values (i.e. pleiotropic), irrespective of the other p-value’s magnitude. This behavior becomes more apparent near or below the genome-wide significance value of 5·10^−8^ (**Fig. 1a**, white line in each plot): even if the other p-value equals 1, FCT and CPMA still assign the SNP to be pleiotropic. By design, composite methods like SumRank and PLACO are resistant to this behavior and require evidence of association in both traits before assigning pleiotropy. Notably, even composite methods show subtle differences in how resistant they are. For example, PLACO’s cross-trait p-value does decrease slowly as one of the p-values becomes increasingly lower, while SumRank plateaus, reflecting a more conservative approach.

**Figure 1.**
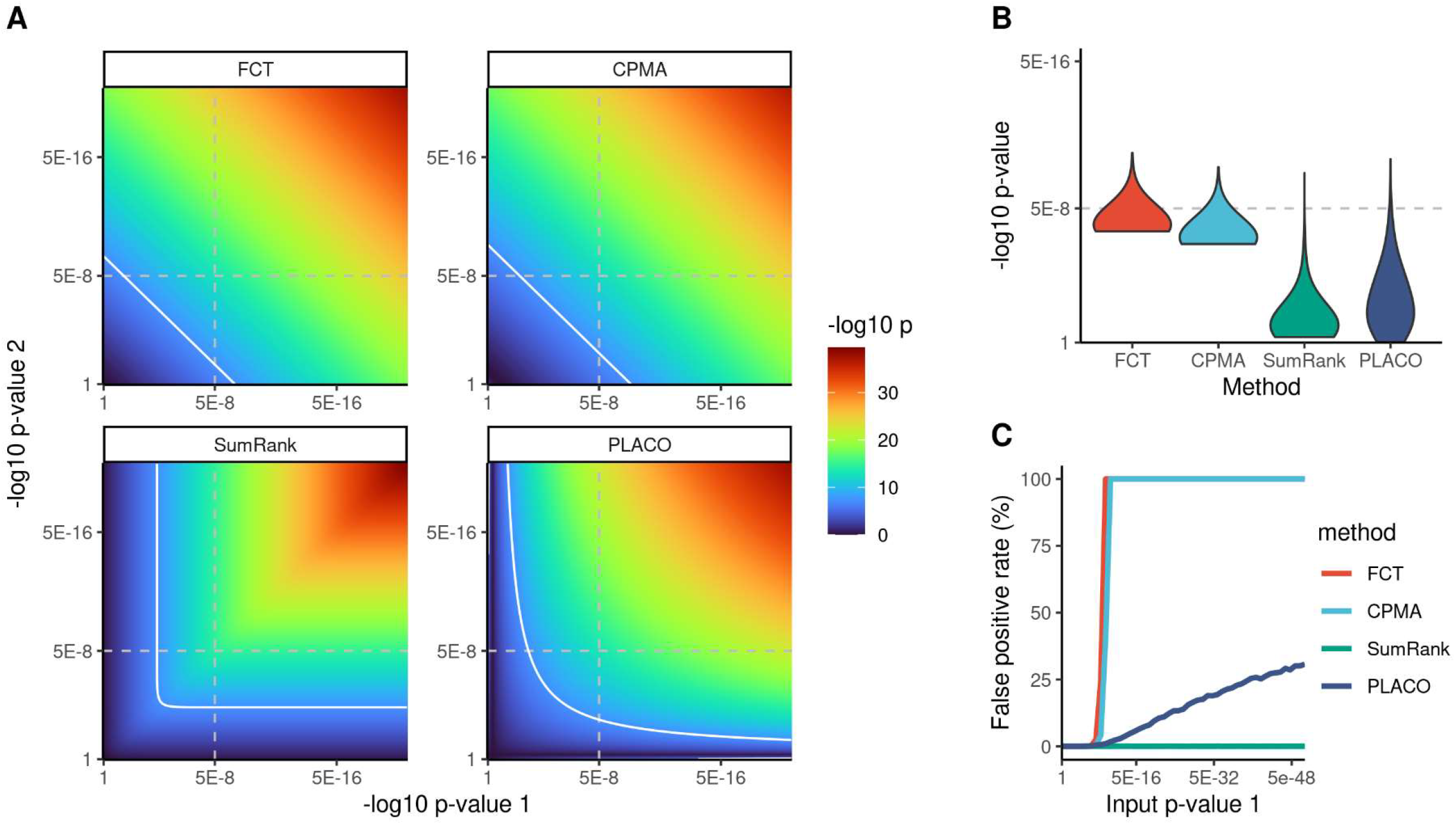
Numerical simulations for two traits. (A) Rejection plots for FCT, CPMA, SumRank, and PLACO. The gray dotted lines denote 5·10^−8^ for the input GWAS p-values, and the white line denotes where the methods output a p-value of 5·10^−8^ or lower. (B) Violin plot of the cross-trait p-values when testing pairs consisting of a p-value of 5·10^−8^ and a uniformly sampled p-value. The y-axis indicates the p-value derived from the methods. (C) Similar to panel B, but the fixed p-value is varied across a range (x-axis) and plotted against the proportion of tests that yield statistically significant results (y-axis).

The implications of the rejection regions were further explored by simulating pairs of p-values. We created one million pairs of p-values drawn from a uniform distribution, representing null conditions. In this scenario, none of the methods had an inflated false positive rate for the two traits (**Supplementary Fig. 1**). Next, we created one million pairs where the first p-value was always set to 5·10^−8^ (i.e., genome-wide significance) and the second p-value was drawn from the uniform distribution. This represents the case where a SNP is only associated with the first trait and therefore should not be classified as pleiotropic. We classified any cross-trait association with a p-value below 5·10^−8^ identified by any method as a false positive finding. Under these conditions, FCT and CPMA had false positive rates of 4.81% and 0.80%, respectively, compared to 0.03% for SumRank and PLACO (**Fig. 1b**). Finally, we investigated the behavior of false positive rates as the fixed p-value threshold was progressively reduced, reaching as low as 5·10^−300^. (**Fig. 1c**). The false positive rates for FCT and CPMA reached 100% if the first p-value was set to 2.34·10^−9^ and 4.07·10^−10^, respectively. PLACO exhibited a more conservative approach yet still resulted in a false positive rate of 25% when the first p-value was set to 1·10^−40^, and 50% when set to 1·10^−111^. In contrast, the false positive rate of SumRank remained 0.03% even when the first p-value was set to 5·10^−300^.

### Genotype simulations for two traits

To emulate a real-world scenario, we conducted a series of population-based genotype simulations with varying genetic architectures. We used UK Biobank genotype data to simulate two correlated synthetic traits, for which we then defined the underlying genetic architectures. Independent SNPs with a minor allele frequency above 5% were selected and a random subset was assigned non-pleiotropic and pleiotropic causal effects, effectively creating hypothetical traits. For the primary simulations, the causal effects were drawn from a multivariate normal distribution. We obtained summary statistics by conducting GWASs, which served as the input for all included cross-trait association methods (**Table 1**). For each method, we clumped the results to obtain lead and correlated SNPs in LD. A detailed description of the procedure can be found in the **Methods** section.

For each of twenty simulations, we generated two traits for two independent cohorts of 100,000 individuals from the UK Biobank. As shown in **Fig. 2a**, each trait had 1000 non-pleiotropic causal SNPs unique to the traits (red) and 1000 pleiotropic causal SNPs shared between them (green). To evaluate the methods, we focused on the number of true pleiotropic SNPs that were detected and the true pleiotropy rate, defined as the proportion of detected SNPs for which the true effect was pleiotropic. The methods varied greatly in performance. For example, the two methods identifying most pleiotropic SNPs were FCT (n = 346) and CPMA (n = 325) (**Fig. 2b**), but these methods also had the lowest true pleiotropy rates (FCT = 47.8%, CPMA = 48.2%) (**Fig. 2c**). In contrast, methods that relied on composite hypotheses generally identified fewer pleiotropic SNPs (e.g., n_conjFDR_ = 246, n_GPA_ = 229, n_SumRank_ = 190), but their true pleiotropy rates were much higher (conjFDR = 98.0%, GPA = 98.5%, SumRank = 99.4%).

**Figure 2.**
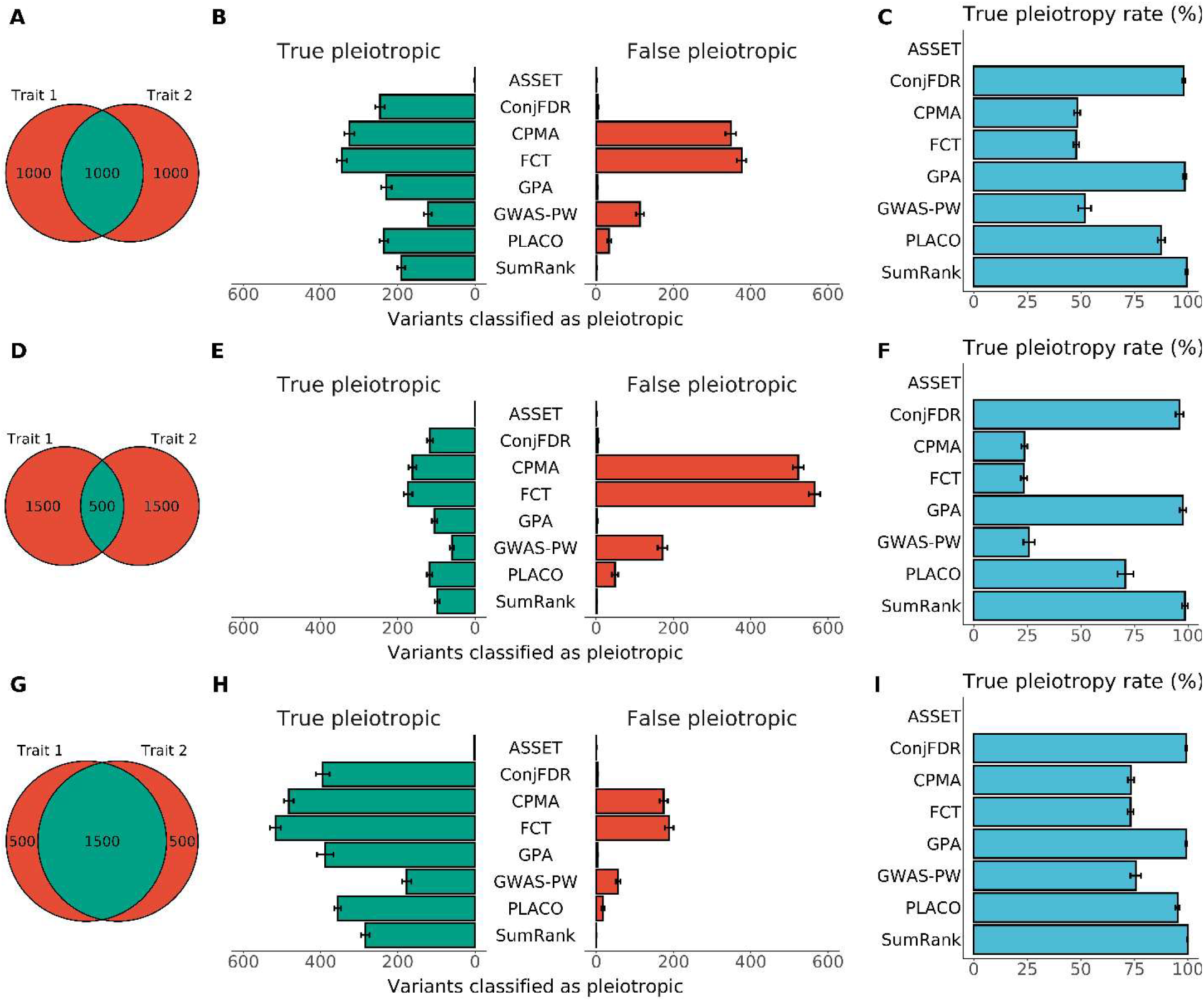
Genotype simulations for two traits. (A, D, G) Schematic of the causal SNPs for the two traits. Green and red indicate pleiotropic and non-pleiotropic SNPs, respectively. (B, E, H) The left columns display the number of shared causal SNPs (green) and unique causal SNPs (red) for various simulation conditions. The right column contains bar plots of the SNPs that each method classified as pleiotropic, and therefore an overview of the number of correct and incorrect findings. (C, F, I) The true pleiotropy rate per method, calculated as the proportion of true pleiotropic SNPs out of the SNPs detected by each method.

GPA and conjFDR inherently correct the false discovery rate in their models. When we applied the Benjamini-Hochberg procedure to the SumRank results, it identified 277 pleiotropic SNPs with a true pleiotropy rate of 96.4%. Crucially, all SNPs discovered by the unadjusted SumRank were also found by conjFDR, which were in turn found by GPA, and all of these were contained within the Benjamini-Hochberg-adjusted SumRank results, indicating a nested pattern of pleiotropic hits across methods (**Supplementary Fig. 2**).

Two methods involving composite hypotheses performed poorly in the two-trait simulations: GWAS-PW, which exhibited a true pleiotropy rate of roughly 50% regardless of the threshold applied to the posterior probability (**Supplementary Fig. 3**); and ASSET, which barely detected any pleiotropic SNPs under these conditions (n = 1).

Overall, most true pleiotropic variants were found by FCT and CPMA while conjFDR, SumRank, and GPA had the highest true pleiotropy rates. This relative pattern of performance remained similar when varying the degree of overlap between the traits (**Fig. 2d-i**), the heritability of the traits (**Supplementary Fig. 4**), the correlation between the effect sizes within each pleiotropic SNP (**Supplementary Fig. 5**), and the sample size of the GWASs (**Supplementary Fig. 6**).

### Numerical simulations for more than two traits

Next, we assessed performance in numerical and genotype simulations with more than two traits. For these simulations, we modified the FCT, CPMA, and SumRank methods to utilize the same subset-based approach as the ASSET method. In brief, given a set of *m* traits, ASSET identifies the subset of traits that is the most likely cross-trait association for a SNP g_i_. Other methods can be modified to similarly detect a subset of traits by testing every subset of traits *k* ∈ *m* and selecting the subset *k* with the lowest p-value that reaches statistical significance. Finally, ASSET also applies a p-value filter: in the analysis of a given SNP g_i_ only traits with a p-value below 0.05 are included. This drastically reduces the number of possible subsets to test, which reduces the false positive rate.

We first performed numerical simulations for the subset-based versions of ASSET, FCT, CPMA, and SumRank. For each SNP, we simulated 10 to 300 trait p-values in total, and the SNPs were causal for 0 to 5 of those traits (see **Methods** section). In these models, we assessed how often the methods classified SNPs to be causal for the non-associated traits (<u>false positive</u>), the truly associated traits (<u>true positive</u>), or only a subset of the truly associated traits (<u>partial positive</u>). First, to illustrate the effect of the p-value filter, we did not apply any filter to the methods, as is typically done in all methods but ASSET. The results showed a striking pattern: in the extreme scenario where 300 traits were analyzed, the false positive rates of all methods varied between 30% and 100% (**Fig. 3a, Supplementary Fig. 7**). We then applied various p-value thresholds to these methods, which resulted in false positive rates that were near 0% for all methods except for CPMA (**Fig. 3b**).

**Figure 3.**
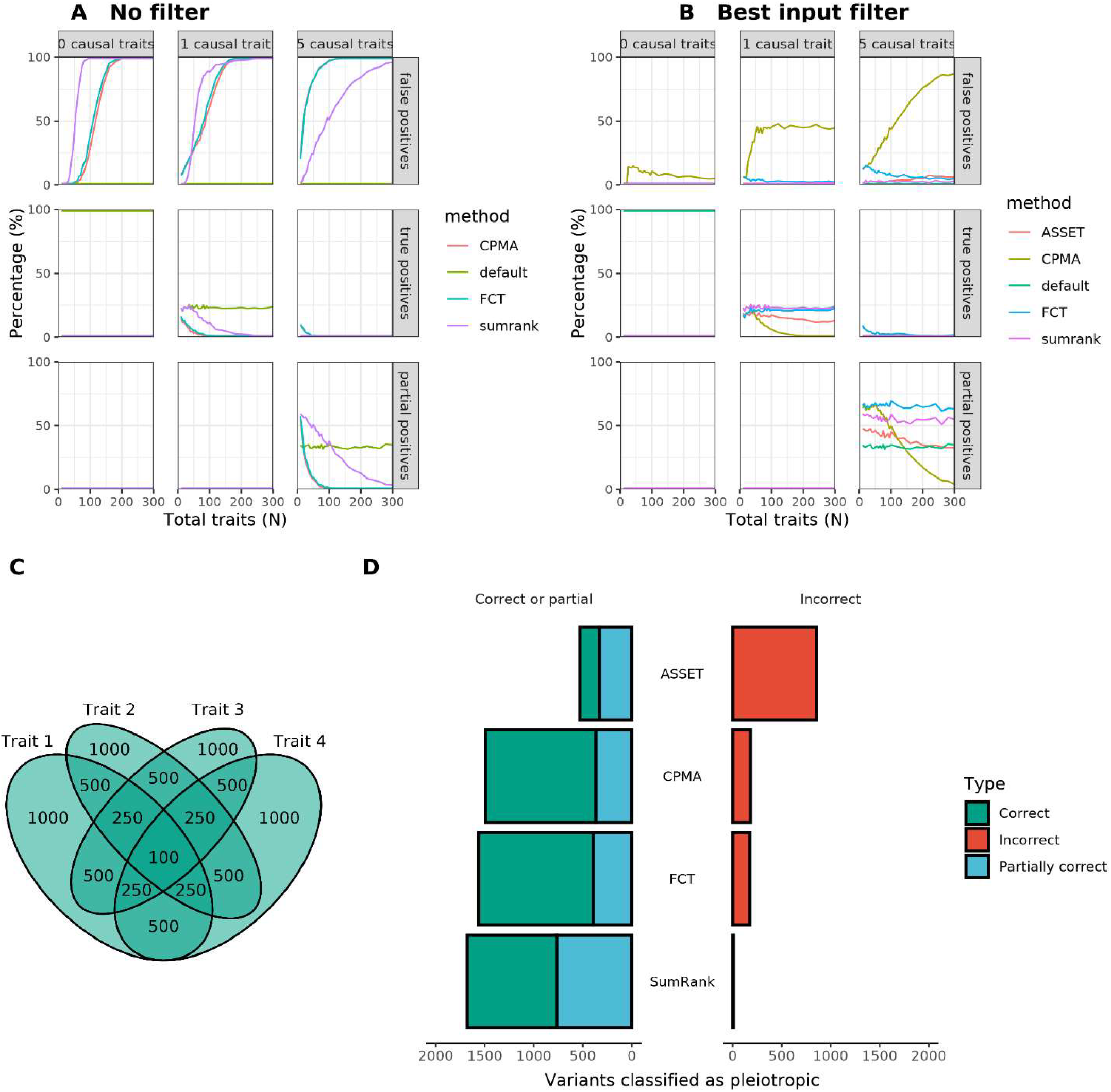
Numerical and genotype simulations for more than two traits. (A) Numerical simulations for SNPs that affect 0, 1, or 5 traits (columns) with metrics of false positives, true positives, and partial positive rates (rows), in analyses of 10 to 300 traits (x-axis). The partial positive rate denotes when a subset of the causal traits was identified, but not all. (B) When only those traits are included that have sufficiently low p-values (see Supplementary Figure 6). (C) Schematic of the causal SNPs for the genotype simulations of four traits. (D) Results from the genotype simulations.

The methods differed in the rates of true positive and partial positive cross-trait associations. For example, for SNPs that were causal for five traits, SumRank had a partial positive rate of 60%, compared to 45% for ASSET. These findings show that ASSET works well, and that other cross-trait association methods can be modified to explore numerous traits simultaneously by implementing a subset-based approach and filtering input p-values.

### Genotype simulations for more than two traits

Next, we repeated the genotype simulations with UK Biobank data. As we limited ourselves to non-overlapping GWASs of 100,000 individuals, we simulated a scenario for four traits, for ASSET as well as the subset-based versions of FCT, CPMA, and SumRank. Each possible combination of traits had causal SNPs, with SNPs being associated with one (n = 1000 per trait), two (n = 500 per combination), three (n = 250), or four traits (n = 100; **Fig. 3c**). The analyses yielded three types of outcomes: pleiotropic SNPs for which all associated traits were detected (<u>true positive</u>), pleiotropic SNPs for which only a subset of associated traits were detected (<u>partial positive</u>), and SNPs for which at least one non-associated trait was detected (<u>false positive</u>). When considering only true positive SNPs, FCT identified most (1169 SNPs), followed by CPMA (1127), ASSET (954), and SumRank (915) (**Fig. 3d**). However, SumRank identified a much larger number of partial positive SNPs (763) compared to FCT (393), CPMA (367), and ASSET (562). Furthermore, the false positive rate for SumRank was only 0.5%, compared to 10.0% for FCT, 10.9% for CPMA, and 1.5% for ASSET.

### Application to psychiatric traits

To further explore the differences between the methods in real-world data, we applied SumRank, FCT, CPMA, and ASSET to the summary statistics for publicly available GWASs of eight prominent psychiatric disorders (**Supplementary Table 1**): anorexia nervosa, attention-deficit/hyperactivity disorder, autism spectrum disorder, bipolar disorder, major depressive disorder, obsessive-compulsive disorder, schizophrenia, and Tourette syndrome. The methods were applied to find the optimal subset of traits that were associated with each SNP.

The original eight GWASs identified 304 loci. Across the four cross-trait methods, 658 novel cross-trait loci were identified. SumRank identified the most novel cross-trait loci (n = 565, **Fig. 4a**), followed by FCT (n = 400), CPMA (n = 244), and finally ASSET (n = 184) (**Supplementary Fig. 8-10**). Of the 658 novel loci, 253 were unique to SumRank, 67 to FCT, and 43 to ASSET (**Fig. 4b**), and 128 loci were found by all four methods. On average, the locus length of the novel loci were similar across the methods (**Supplementary Fig. 11**).

**Figure 4.**
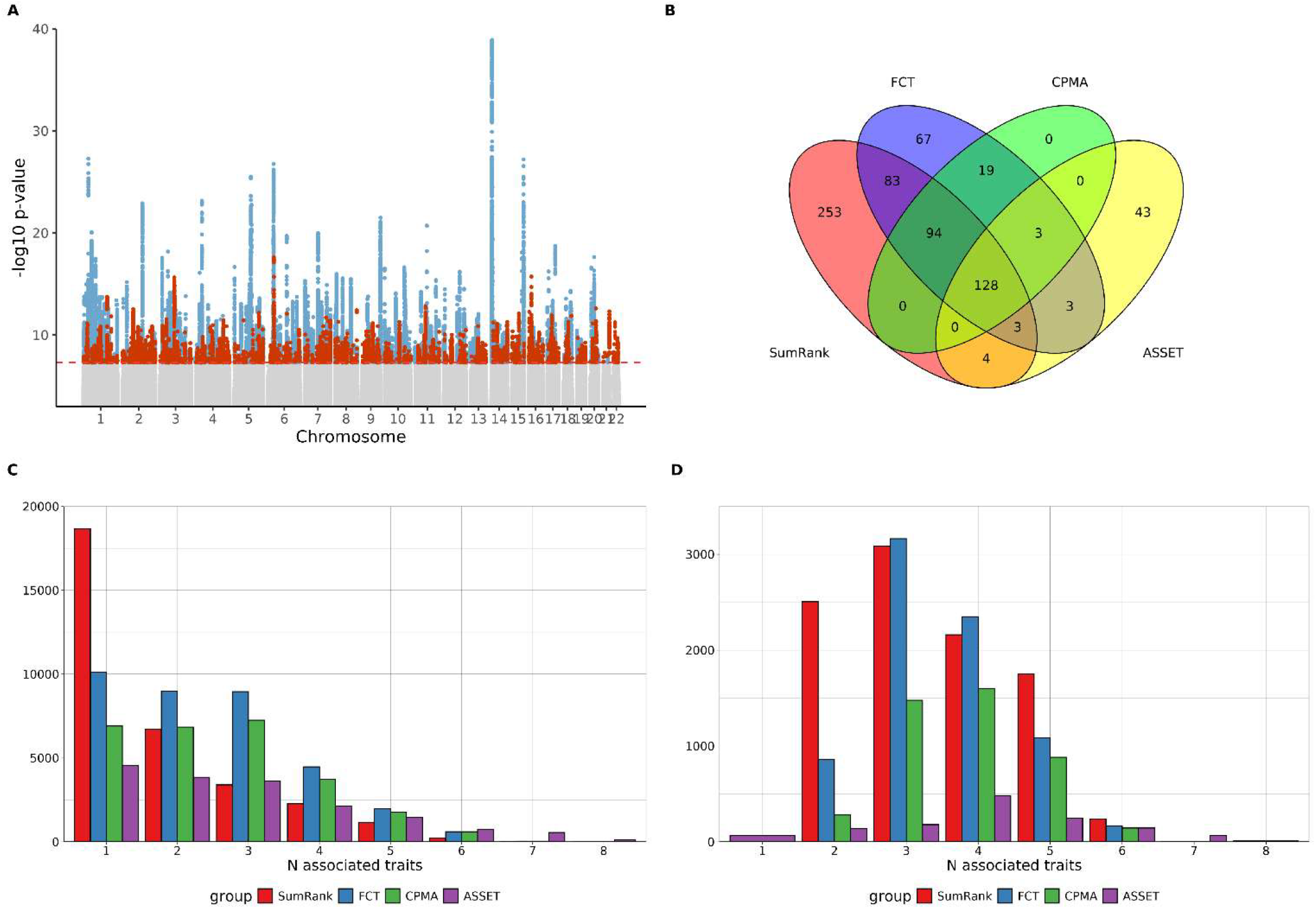
Cross-trait associations of eight psychiatric disorders. (A) Manhattan plot showing the results for the SumRank analysis. Each point represents the p-value for the optimal subset for a given SNP. Blue SNPs are genome-wide significant SNPs in loci that had previously been identified in one of the original GWASs. Dark orange SNPs represent genome-wide significant SNPs in novel loci. (B) Venn diagram of the overlap in detected regions by SumRank, FCT, CPMA, and ASSET. (C) Number of associated traits for the detected SNPs within the regions that were genome-wide significant in at least one of the original psychiatry GWASs. (D) Number of associated traits for the detected SNPs in novel loci.

Next, we considered the distribution of associated traits that each method found in the 304 previously identified loci. SNPs in most of these loci were only associated with one of the original GWASs. SumRank found a similar pattern, where the majority of identified SNPs in these loci were still associated with only one trait (**Fig. 4c**). In contrast, FCT and CPMA assigned the majority of these SNPs to be pleiotropic across two or three traits. This suggests that FCT and CPMA are more likely to assign SNPs as pleiotropic when the SNP is genome-wide significantly associated with one trait. This confirms the rejection plots, and the higher rate of false positives observed in our previous simulations (see **Fig. 1a** and **Fig. 3b**).

When considering the newly discovered loci, the distribution of associated traits was similar across the different methods (**Fig. 4d**). Interestingly, the unique loci that SumRank identified were primarily due to a relative enrichment of SNPs pleiotropic for two traits. This is in line with the high partial positive rate that SumRank had in the simulations (see **Fig. 3d**), suggesting that SumRank finds more true positive pleiotropic loci than other methods, although it will not identify all the traits associated with these loci.

To further explore any downstream differences between the methods, we functionally annotated the SNPs. As expected from previous literature, the SNPs that were implicated by the cross-trait association methods were most likely to be intronic or intergenic (**Supplementary Fig. 12**). For each method, as the number of associated traits increased, the SNPs were increasingly likely to be located in introns. When considering tissue-specific gene enrichment for the mapped genes, the results of all methods pointed towards enrichment in brain-related tissues from the GTEx v8 atlas (**Supplementary Fig. 13**).

## Discussion

The evaluation of summary-level cross-trait association methods revealed distinct differences in their performance but also common limitations. First, in numeric and genotype simulations of two traits we observed substantial inflation of false positive rates for various methods, warranting careful review of previous work that has been published with these methods. The best performing methods–conjFDR, SumRank, and GPA–all achieved high true positive rates; meanwhile, time the methods strictly controlled the false positive rate by using a composite null hypothesis. Second, in numeric and genotype simulations of up to 300 traits, we showed that multiple methods can achieve reasonable power and control the false positive rate by adapting the subset approach introduced by ASSET. Amongst these adapted methods, SumRank had the highest true positive rate while maintaining a stringent false positive rate, closely followed by ASSET. Third, our application to psychiatric traits reaffirmed that cross-trait association methods can drastically improve the yield from GWASs by leveraging pleiotropy.

The widespread interest in pleiotropy and the accessibility of GWASs have led to studies focusing on cross-trait associations across tens^7,10-13^, hundreds^1,14,15^, or even thousands of traits^16-18^. Overall, three different approaches should be distinguished to study cross-trait associations across more than two traits. The first and most stringent approach identifies SNPs as pleiotropic only if they reach genome-wide significance in all underlying GWASs^1^. While this method minimizes false positive findings, it risks many false negative cross-trait associations due to loci with weak signals. The second approach is to analyze all traits pairwise. This approach successfully revealed robust cross-trait loci in previous studies^7^. However, being an extension of the first approach, it underestimates loci with more than two associated traits, especially if the signal is weak. Our findings indicate that a third approach, examining every subset of possible traits^4^, is most effective if more than two traits are studied. We show that this approach, which was first implemented in ASSET, can be effectively applied to other methods when combined with p-value filtering approaches to mitigate severe inflation of false positives. Overall, our findings suggest that two cross-trait association methods, SumRank and ASSET, can both be used to validly study hundreds of traits simultaneously.

Regardless of the number of traits, our study showed significant variation in false positives rates of different methods. Some methods provide strict control (e.g., SumRank), others lenient control (e.g., PLACO), while some fail to control false positives adequately (e.g., non-composite hypothesis methods). Even among methods with seemingly appropriate control, false positives were inflated under specific conditions. For example, composite hypothesis methods like PLACO can produce severe inflation of false positives if a SNP associates strongly with one trait but not the other, a limitation noted by PLACO’s authors^8^. Still, no singular method objectively outperformed all other methods. SumRank, for example, performed well across all simulation conditions, but the choice to use other methods can be justified depending on the characteristics of the study and the relative importance ascribed to false positive and false negative rates. Stringent methods like SumRank may be best suited for functional follow-up studies requiring precise SNP prioritization, while more lenient methods may be suitable for explorative studies of less well-powered GWASs. Given the centrality of pleiotropy in human health, there is an urgent need for consensus on the minimal requirements to properly test and validate cross-trait methods, standardization of benchmark simulations and ground truth datasets, and establishment of metrics by which these methods should be evaluated.

The results should be interpreted with several limitations in mind. First, while our simulations cover a broad spectrum of scenarios, they cannot capture all the complexities of real-world situations. For example, we selected causal SNPs that were reasonably independent, whereas genomic regions typically harbor multiple causal SNPs. Second, we focused on cross-trait associations rather than pleiotropy itself. Cross-trait association methods cannot distinguish horizontal from vertical pleiotropy. Thus, results from these methods should not be interpreted as causal without appropriate follow-up work using pleiotropy-specific methods and functional validation studies. Third, we only evaluated cross-trait association methods that are commonly used or cited.

This study elucidates the intricacies of cross-trait genetic association methods, highlighting significant shortcomings and strengths of various approaches. Our work emphasizes the need for more reliable cross-trait association methods and a standardized simulation framework to validate and develop them. By advancing such a framework and by introducing SumRank, we hope to stimulate methodological innovation in the study of cross-trait associations and pleiotropy.

## Methods

### SumRank

The SumRank method is based on the idea that pleiotropy can be assessed from GWAS summary statistics by ranking the p-values from the smallest (first rank) to the largest (maximum rank) and subsequently summing the ranks per SNP. Considering *m* traits with *N* SNPs, ***P*** represents an *N* x *m* matrix, where each column contains the ranks of the p-values within each trait. The null hypothesis tested with the SumRank method represents the scenario where at most m - 1 traits are associated with the SNP. Under the null hypothesis, the distribution of ranks within a GWAS is uniform, and therefore the probability that rank *P*_*i,j*_ (SNP *i* and trait *j*) is equal to any rank is 1/N. The ranks can subsequently be summed per SNP to create ***R***, which is a vector of length *N* with *m* ≤ *R*_*i*_ ≤ *mN*.

SumRank tests the probability of ***R***_***i***_ being equal to rank *r* or more extreme under a null hypothesis of randomly assigned ranks. Consider a set of vectors *x*_*1*_, *…, x*_*m*_ such that *x*_*1*_ *+* … + *x*_*m*_ = ***R***_***sum***,***i***_ with 1 ≤ *x*_*i*_ ≤ ***N***. Let ***S*** be a set of solutions with ***x***_***i***_ ≥ 1 and let ***S***_i_ be the set of solutions with ***x***_***i***_ < ***N*** for exactly that *i*, then:

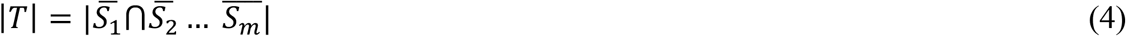

Based on the inclusion/exclusion criteria we can rewrite eq. (3) as:

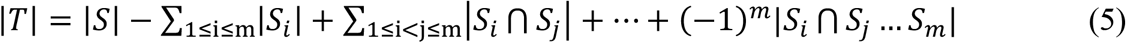

where |***S***| is a combinatorial composition which equals 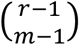, or in other words, how many combinations of *m*-1 positive integer numbers can give us a sum equal to *r* −1. Therefore, |*S*_i_| = 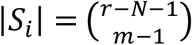, and 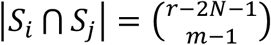, etc., and:

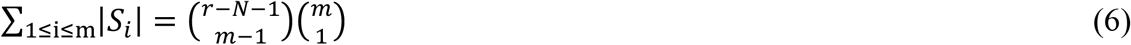

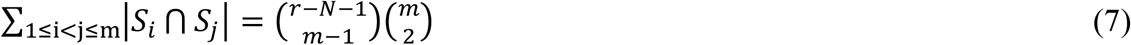

etc. It is now possible to simplify eq. (4):

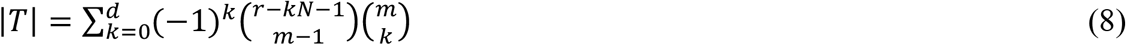

where:

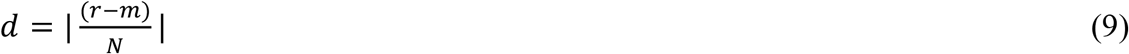

We can further divide |***T***| in eq. (7) by the total number of vectors of length *m* – as they arise with equal probability under the null hypothesis – to derive:

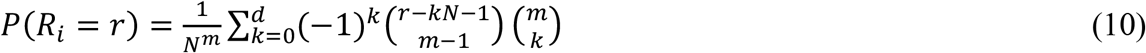

Finally, we compute the p-value for SumRank:

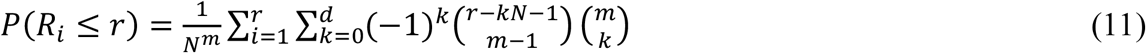

In practice, we want to test the null hypothesis when ***R***_***i***_ is much smaller than ***N***, and thus *d* will always be approximately zero. We can then simplify eq. (11):

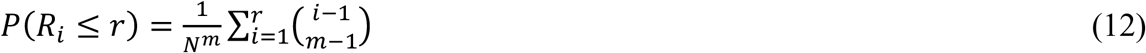

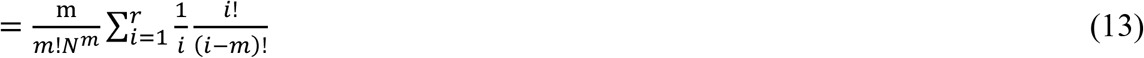

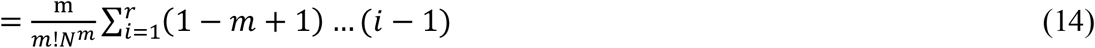

As the number of independent SNPs increases, the formula can be further simplified to eq. (3), shown in the Results section:

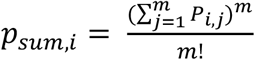

### Genotype data

We used release version 3 for the imputed genotype datasets from the UK Biobank. The design of the UK Biobank resource has been approved by the North West Centre for Research Ethics Committee, and written informed consent was provided by all participants. The procedures for genotyping, imputation, and quality control have been described elsewhere^19^. In brief, blood samples from 488,377 participants were assayed with two different genotyping arrays: the Applied Biosystems UK BiLEVE Axiom Array by Affymetrix and the Applied BioSystems UKB Axiom Array. After quality control of the markers and samples, the autosomes were phased with SHAPEIT3^20^, using the 1000 Genomes phase 3 dataset as a reference^21^, and subsequently imputed against the Haplotype Reference Consortium (HRC) dataset^22^. The data were also separately imputed against a combination of the 1000 Genomes phase 3 and the UK10K datasets^23^, and all data were combined – using the HRC imputation when a SNP was available in both – into a dataset consisting of 93,095,623 autosomal variants in 478,442 individuals.

### Genotype simulations

The genotype simulations were conducted in a series of steps. First, we identified independent SNPs based on the full genetic dataset of the UK Biobank. We excluded individuals with a call rate of < 0.99; and we excluded SNPs with any missingness, a violation of the Hardy-Weinberg equilibrium (p < 10^−6^), and less or more than two alleles. To obtain the independent SNPs we used PLINK2^24^ with --indep-pairwise 50 5 0.05.

Second, we created 100 simulations where we randomly selected a subset of the independent SNPs. For the traits that we aimed to simulate, we assigned each of these SNPs an effect size. In the case of two traits, we selected 1000 causal SNPs that only affected the first trait, 1000 causal SNPs that only affected the second trait, and 1000 causal SNPs that were pleiotropic and thus affected both traits. The effect sizes were drawn from a normal distribution *N*(0, 1), and we set the Pearson’s correlation between the effect sizes within a pleiotropic SNP to 0.5.

Third, for each simulation we selected a subset of 100,000 individuals per trait, and we calculated the simulated traits based on the causal SNPs. The assigned trait value was set as the sum of the genetic component scaled by the heritability, and a complementary non-genetic component:

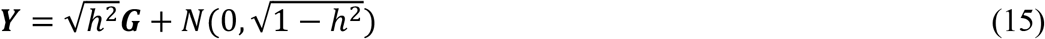

The phenotypic correlation between the traits was forced to 0.5, and the h^2^ for both traits was set to 0.5.

Fourth, we conducted GWASs for the simulated traits and applied the pleiotropy methods. The GWASs were conducted with fastGWA^25^. Each GWAS was run on 100,000 individuals. The samples were non-overlapping, as overlap between the samples can lead to bias in the estimates^26^, and correcting for this bias is beyond the scope of the current study.

Finally, to see whether the methods identified the non-pleiotropic or pleiotropic causal SNPs, we clumped the results using PLINK2^24^ and assessed whether any causal SNPs were in LD with the lead SNPs. We based the LD structure for clumping on the total sample that partook in the simulated GWASs. We set independent SNPs to be those that were genome-wide significant (p < 5·10^−8^) and not in LD after clumping (r^2^ < 0.6). A second round of clumping was performed against r^2^ < 0.1 to define the lead SNPs. For each lead SNP, we checked whether any causal SNPs in LD were pleiotropic or non-pleiotropic.

The simulations relied on several set parameters and modeling decisions. We performed multiple sensitivity analyses to understand how the behavior of the methods changed. For the sample size we reran the simulations with 50,000 and 150,000 individuals per GWAS. For heritability we also simulated h^2^ being equal to 0.1, 0.2, 0.3 and 0.4. Finally, we changed the Pearson’s correlation of the effect sizes within pleiotropic SNPs to 0.0 and 1.0.

### Numerical simulations for more than two traits

To assess the false positive and true positive rates for more than two traits, we simulated associations between causal SNPs and 10 to 100 traits. SNPs were defined as causal for zero, one, or five traits. The effect sizes per minor allele were drawn from a normal distribution N(0, τ) with:

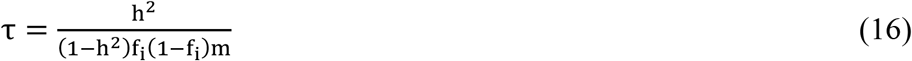

where *h*^*2*^ is the SNP heritability and *m* is the total number of causal SNPs. For our simulations, the SNPs were simulated as having a minor allele frequency of 0.1, an h^2^ of 0.25, and an *m* of 1000. The genotype matrix **G** was generated for 100,000 individuals given the minor allele frequency. Error terms and the traits were calculated as:

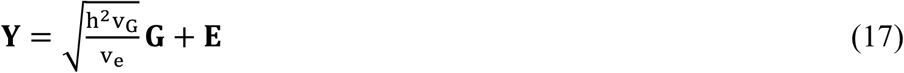

where **E** is a matrix for error terms per person (rows) per trait (columns) drawn from distribution N(0, τ), v_G_ is the genotypic variance per SNP, v_e_ is the column-wise variance of **E**, and 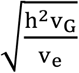 weighs the genetic and phenotypic components. The SNPs in **G** were then associated with 10 to 100 traits using linear regression models, where the non-causal traits were drawn from a normal distribution N(0, 1). The resulting p-values were used as input for the cross-trait association methods.

## Acknowledgements

This research has been conducted using the UK Biobank Resource under application number 23509. This work was supported by the Netherlands Organization for Scientific Research (NWO/ZonMW grant 016.VICI.170.200 [SL, HT], NWO Veni grant 916.19.151 [SL, HA], NOW Veni grant 1936320 [GR]).We thank SURFsara (www.surfsara.nl) for their support in using the Dutch national supercomputer Snellius.

## Data availability

All summary statistics from the PGC can be obtained from https://www.med.unc.edu/pgc/download-results/. The raw UK Biobank data can be obtained through a data access application available at https://www.ukbiobank.ac.uk.

## Code availability

The code for SumRank is available through the *ctar* R package, which can be downloaded at https://github.com/slamballais/ctar.

## References

1. Watanabe, K. et al. A global overview of pleiotropy and genetic architecture in complex traits. Nat Genet 51, 1339–1348 (2019).

2. Visscher, P.M. et al. 10 Years of GWAS Discovery: Biology, Function, and Translation. Am J Hum Genet 101, 5–22 (2017).

3. Cotsapas, C. et al. Pervasive sharing of genetic effects in autoimmune disease. PLoS Genet 7, e1002254 (2011).

4. Bhattacharjee, S. et al. A subset-based approach improves power and interpretation for the combined analysis of genetic association studies of heterogeneous traits. Am J Hum Genet 90, 821–35 (2012).

5. Chung, D., Yang, C., Li, C., Gelernter, J. & Zhao, H. GPA: a statistical approach to prioritizing GWAS results by integrating pleiotropy and annotation. PLoS Genet 10, e1004787 (2014).

6. Andreassen, O.A., Thompson, W.K. & Dale, A.M. Boosting the power of schizophrenia genetics by leveraging new statistical tools. Schizophr Bull 40, 13–7 (2014).

7. Pickrell, J.K. et al. Detection and interpretation of shared genetic influences on 42 human traits. Nat Genet 48, 709–17 (2016).

8. Ray, D. & Chatterjee, N. A powerful method for pleiotropic analysis under composite null hypothesis identifies novel shared loci between Type 2 Diabetes and Prostate Cancer. PLoS Genet 16, e1009218 (2020).

9. Salinas, Y.D., Wang, Z. & DeWan, A.T. Statistical Analysis of Multiple Phenotypes in Genetic Epidemiologic Studies: From Cross-Phenotype Associations to Pleiotropy. Am J Epidemiol 187, 855–863 (2018).

10. Mocci, E. et al. Genome wide association joint analysis reveals 99 risk loci for pain susceptibility and pleiotropic relationships with psychiatric, metabolic, and immunological traits. PLoS Genet 19, e1010977 (2023).

11. Xue, H., Wu, C. & Pan, W. Leveraging existing GWAS summary data of genetically correlated and uncorrelated traits to improve power for a new GWAS. Genet Epidemiol 44, 717–732 (2020).

12. McGrath, I.M., International Endometriosis Genetics, C., Montgomery, G.W. & Mortlock, S. Genomic characterisation of the overlap of endometriosis with 76 comorbidities identifies pleiotropic and causal mechanisms underlying disease risk. Hum Genet 142, 1345–1360 (2023).

13. Rashkin, S.R. et al. Pan-cancer study detects genetic risk variants and shared genetic basis in two large cohorts. Nat Commun 11, 4423 (2020).

14. Li, Y. et al. Combined genome-wide association study of 136 quantitative ear morphology traits in multiple populations reveal 8 novel loci. PLoS Genet 19, e1010786 (2023).

15. Tanha, H.M., Sathyanarayanan, A., International Headache Genetics, C. & Nyholt, D.R. Genetic overlap and causality between blood metabolites and migraine. Am J Hum Genet 108, 2086–2098 (2021).

16. Barrio-Hernandez, I. et al. Network expansion of genetic associations defines a pleiotropy map of human cell biology. Nat Genet 55, 389–398 (2023).

17. Oblong, L.M. et al. Principal and independent genomic components of brain structure and function. Genes Brain Behav 23, e12876 (2024).

18. Zhang, Z. et al. A scalable approach to characterize pleiotropy across thousands of human diseases and complex traits using GWAS summary statistics. Am J Hum Genet 110, 1863–1874 (2023).

19. Bycroft, C. et al. The UK Biobank resource with deep phenotyping and genomic data. Nature 562, 203–209 (2018).

20. O’Connell, J. et al. Haplotype estimation for biobank-scale data sets. Nat Genet 48, 817–20 (2016).

21. 1000 Genomes Project Consortium et al. A global reference for human genetic variation. Nature 526, 68–74 (2015).

22. McCarthy, S. et al. A reference panel of 64,976 haplotypes for genotype imputation. Nat Genet 48, 1279–83 (2016).

23. Huang, J. et al. Improved imputation of low-frequency and rare variants using the UK10K haplotype reference panel. Nat Commun 6, 8111 (2015).

24. Chang, C.C. et al. Second-generation PLINK: rising to the challenge of larger and richer datasets. Gigascience 4, 7 (2015).

25. Jiang, L. et al. A resource-efficient tool for mixed model association analysis of large-scale data. Nat Genet 51, 1749–1755 (2019).

26. LeBlanc, M. et al. A correction for sample overlap in genome-wide association studies in a polygenic pleiotropy-informed framework. BMC Genomics 19, 494 (2018).

27. Fisher, R.A. Statistical Methods for Research Workers, (Oliver and Boyd, Edinburgh, 1925).

