## Supplementary Figures for "A comprehensive benchmarking and validation study of cross-trait association methods"

**Supplementary Figure 1. Q-Q plots for FCT, CPMA, SumRank, and PLACO given uniformly sampled p-values for two phenotypes.** CPMA = Cross-Phenotype Meta-Analysis; FCT = Fisher's Combination Test; PLACO = *Pleiotropic Analysis under COmposite null hypothesis*.

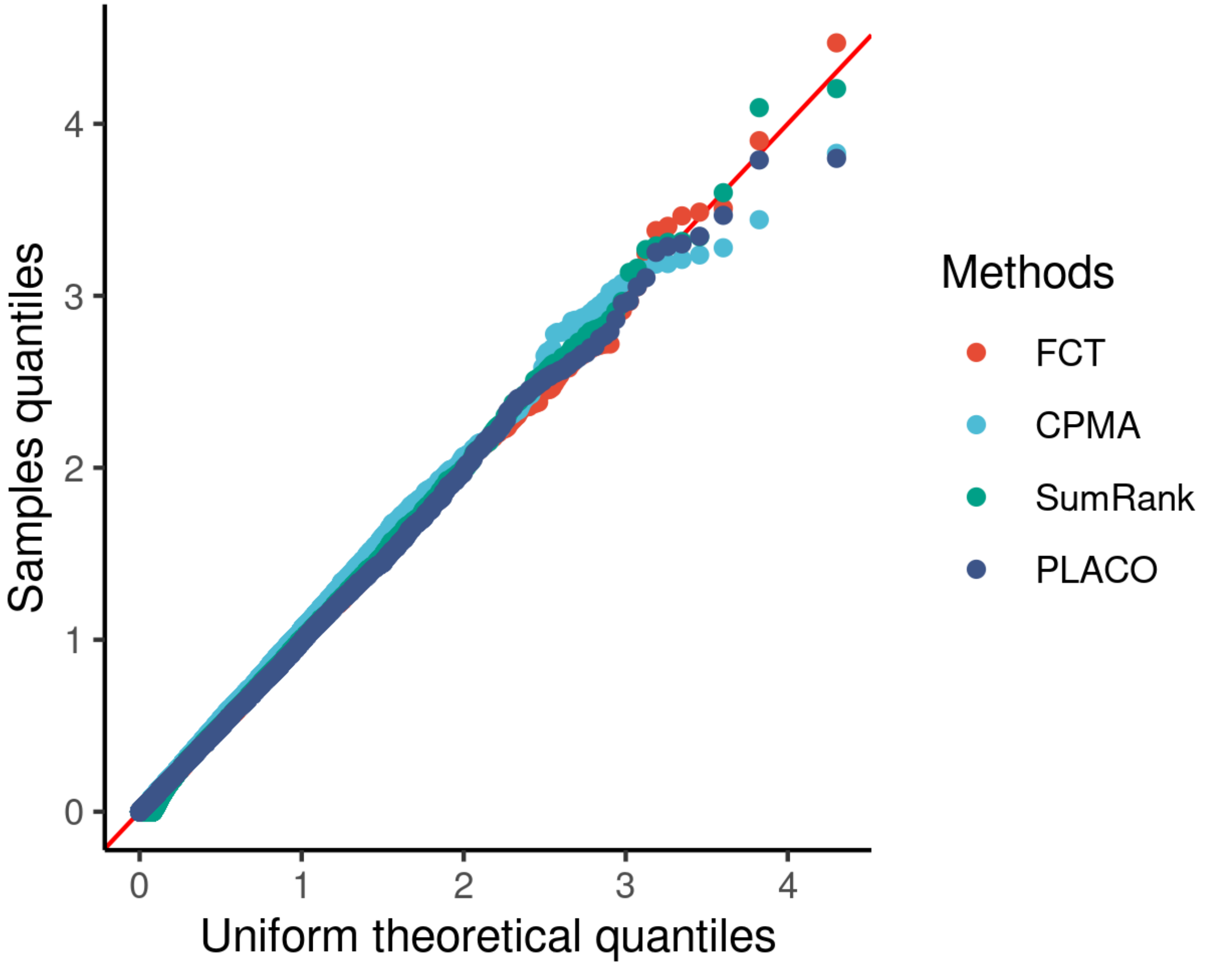

**Supplementary Figure 2. Venn diagram of pleiotropic SNPs identified by SumRank, conjFDR, GPA, and Benjamini-Hochberg-adjusted SumRank**.

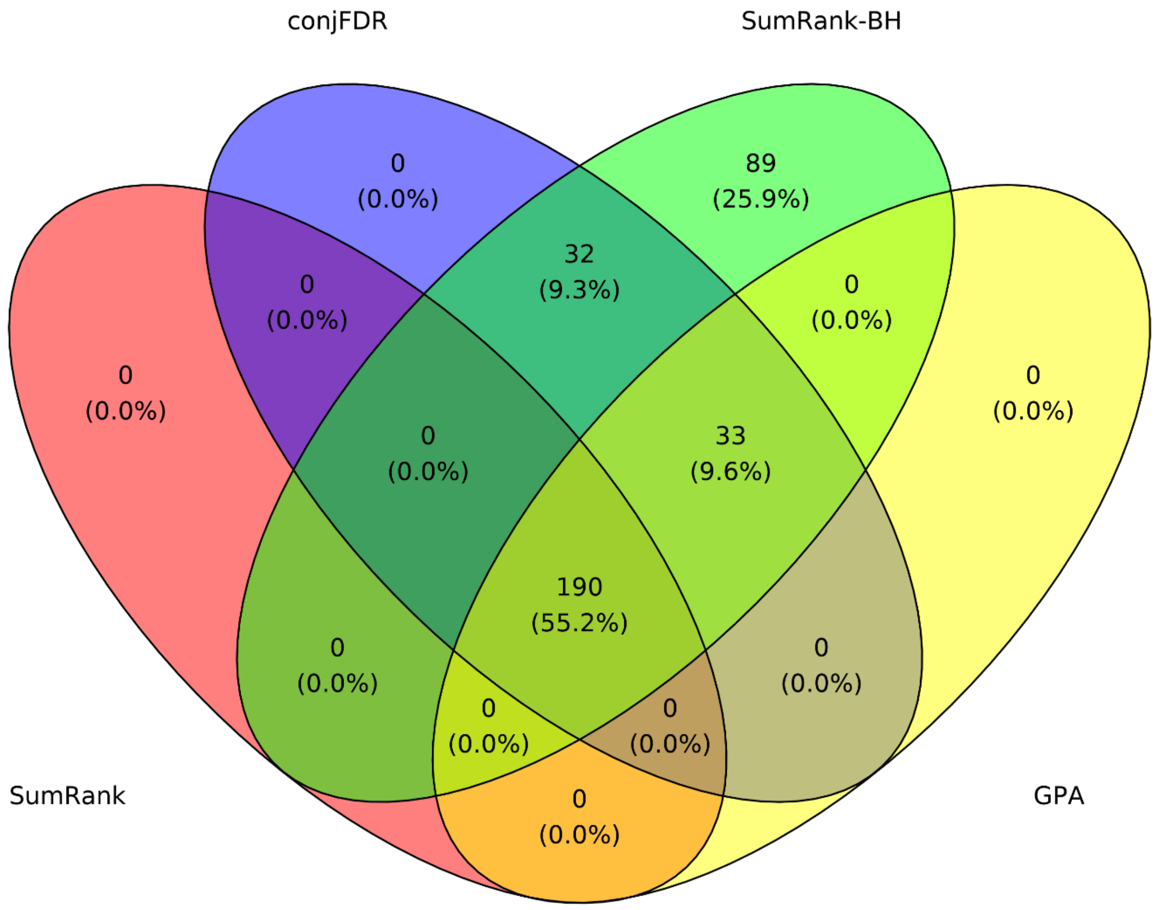

**Supplementary Figure 3. Simulation results for GWAS-PW given varying decision thresholds**. The ribbons represent one standard deviation from the means. PP = posterior probability.

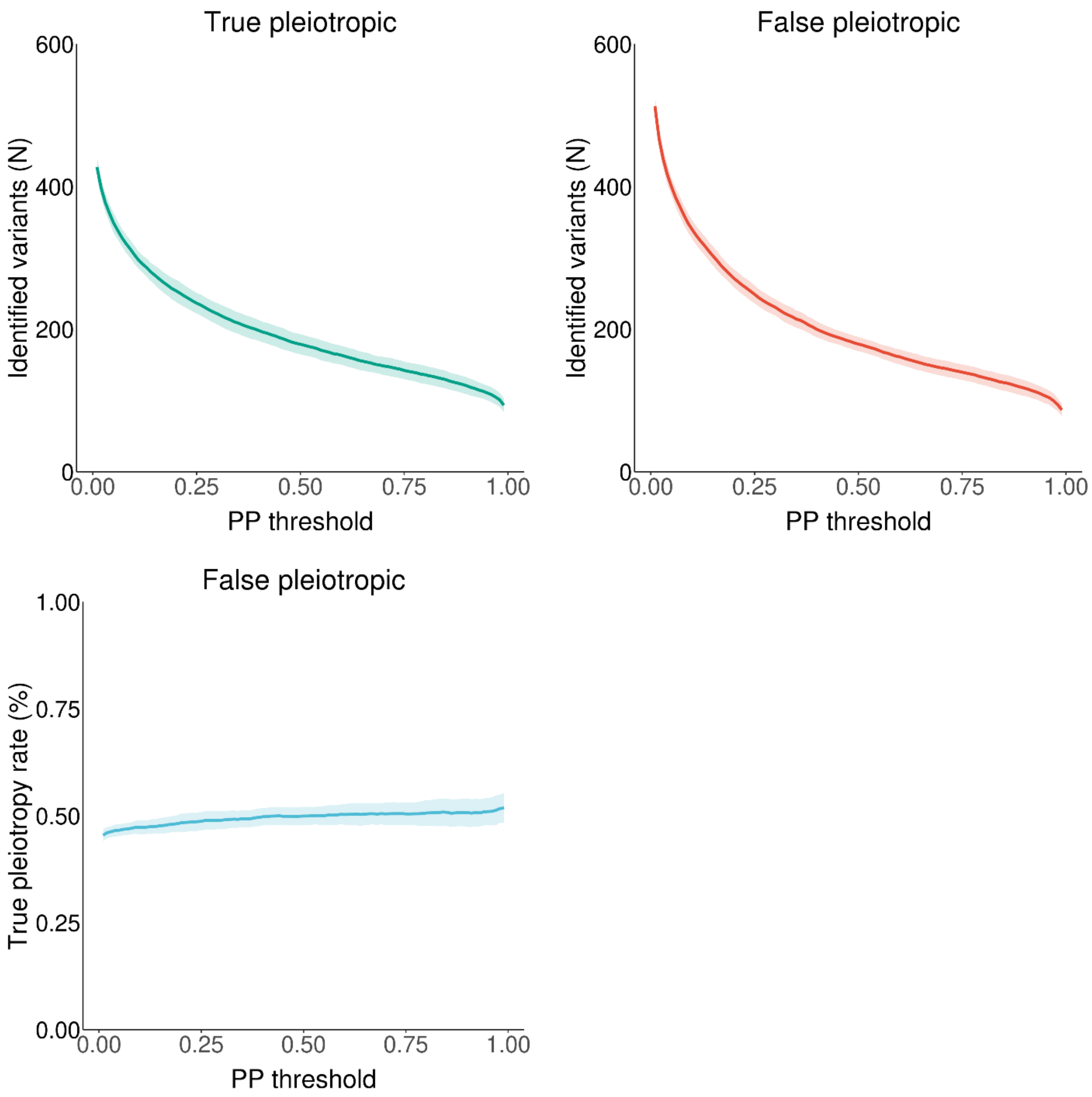

**Supplementary Figure 4. Results for the genotype simulations of two traits, varying the heritability of both traits ($h^2$ = 0.1, 0.2, 0.3, 0.4).**

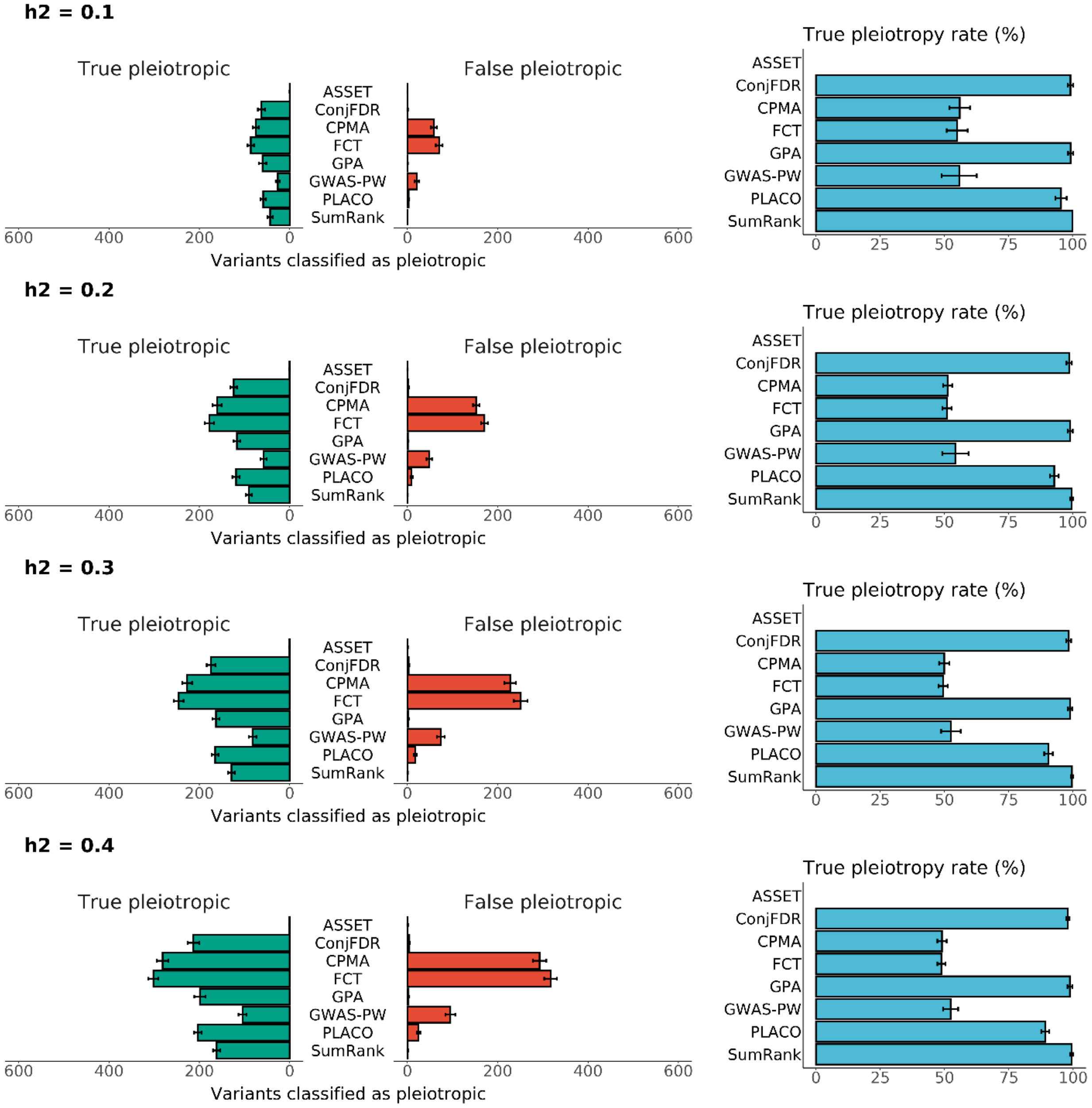

**Supplementary Figure 5. Results for the genotype simulations of two traits, varying the correlation between the effect sizes within a pleiotropic SNP (r = 0.0, 1.0).**

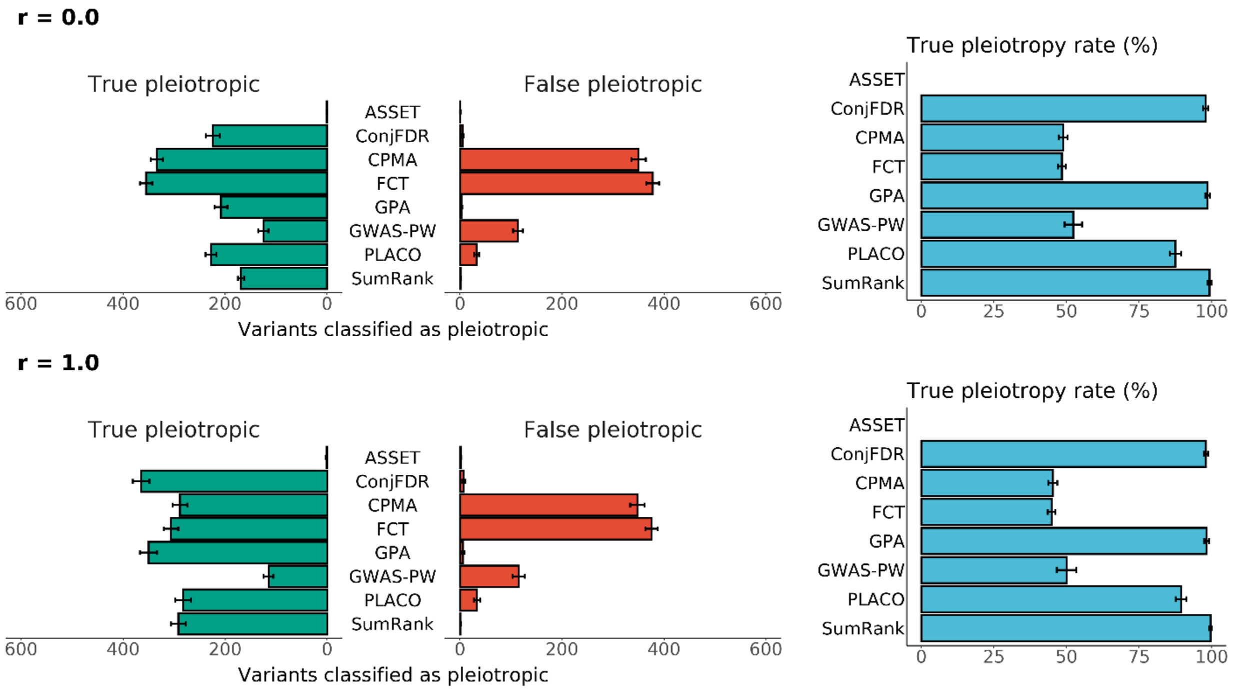

**Supplementary Figure 6. Results for the genotype simulations of two traits, varying the sample size of the input GWASs (N = 50,000, 150,000).**

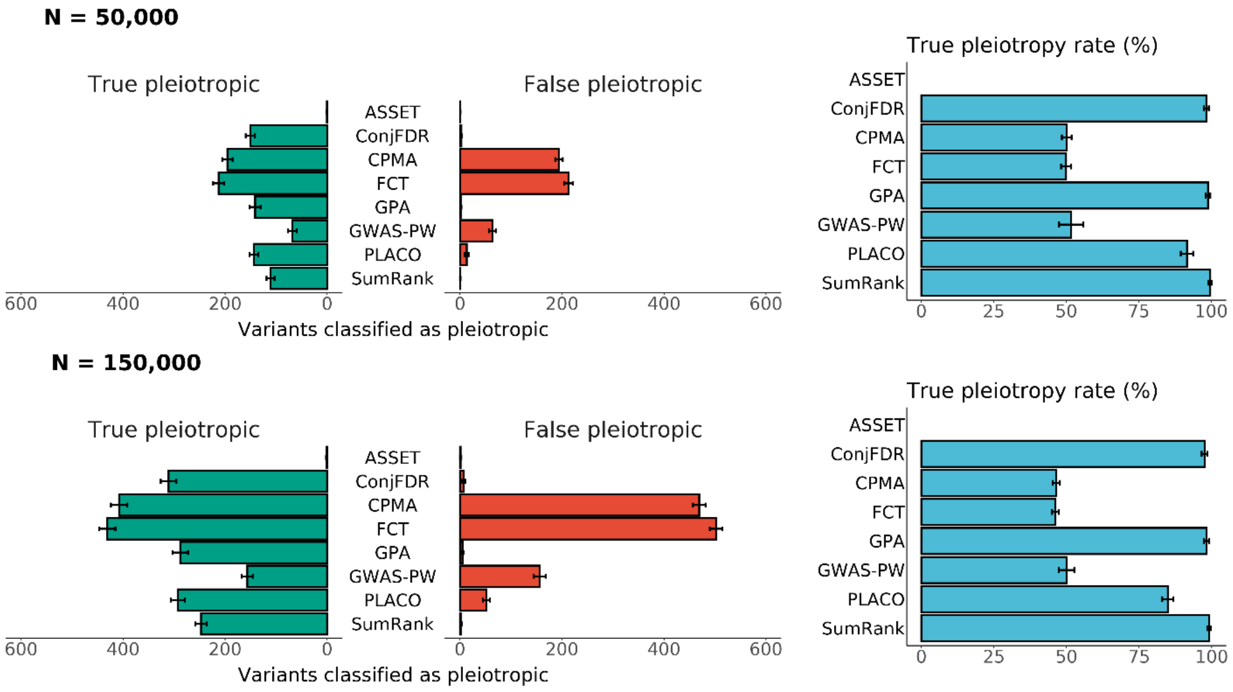

**Supplementary Figure 7.** Numerical simulations for SNPs that affect (A) 0 or (B) 5 traits with metrics of false positives, true positives, and partial positives (rows), with various p-value filtering methods (columns), in analyses of 10 to 300 traits (x-axis). The partial positive rate denotes when a subset of the causal traits were identified, but not all. The p-values were filtered at 1 (no filter), 0.05, [1/*m*], [1/(*m*·log(*m*))], or [1/(m·sqrt(*m*))], where *m* is the number of traits in the analysis.

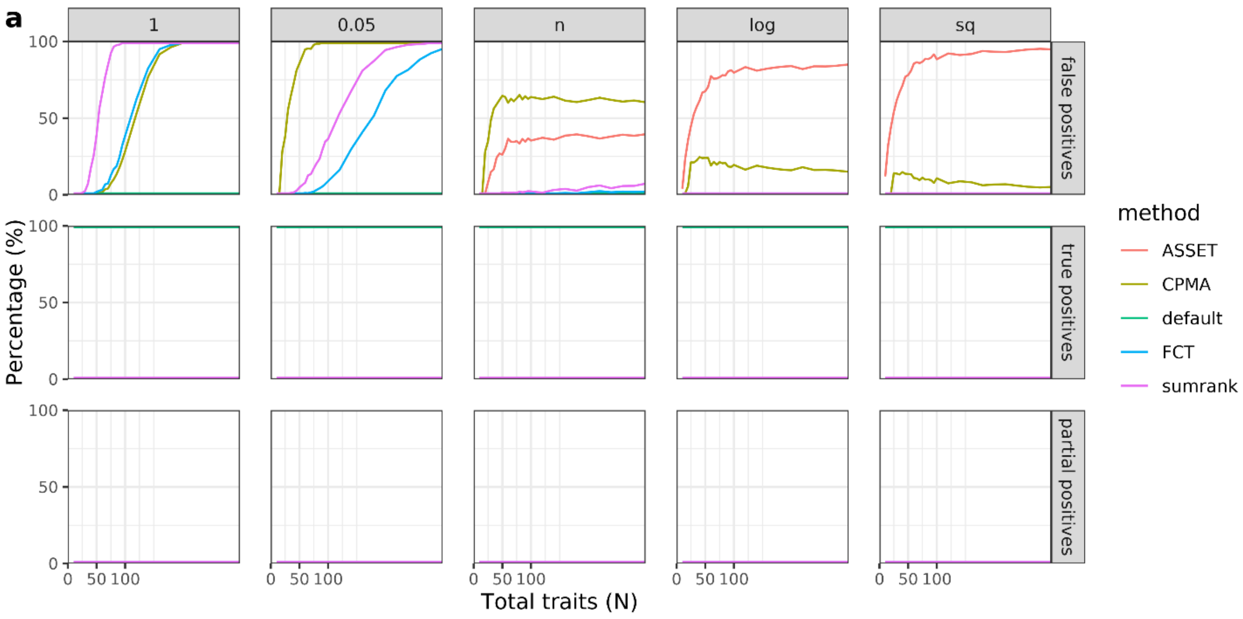


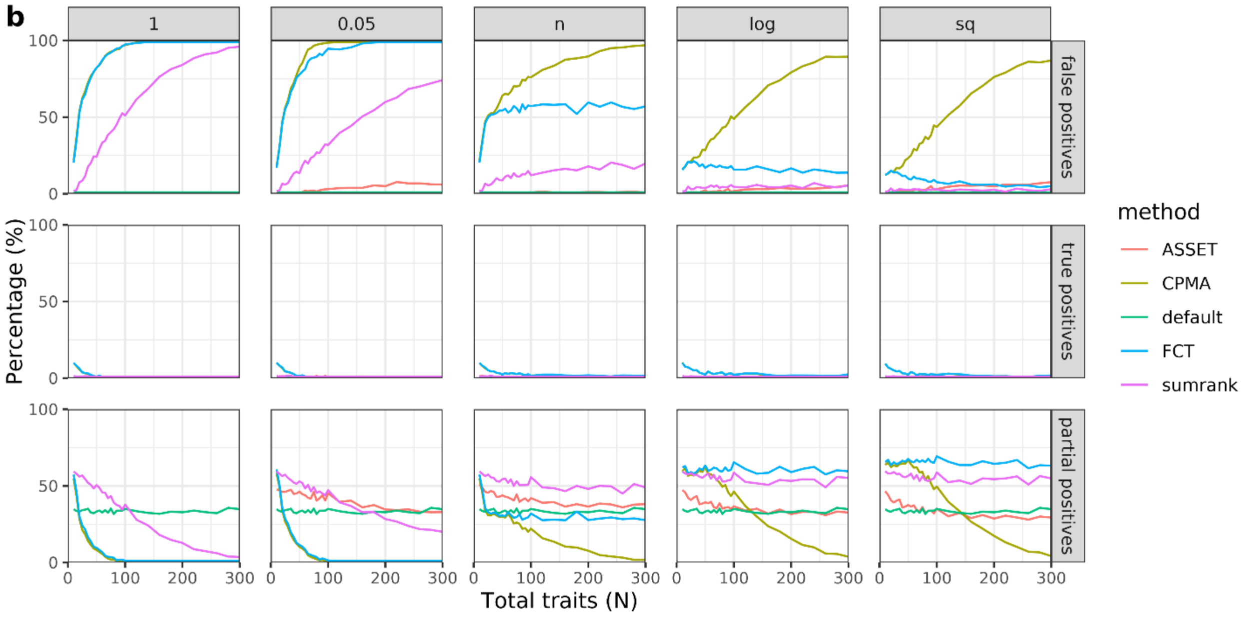

**Supplementary Figure 8. A Manhattan plot representing the FCT results for cross-trait associations across psychiatric traits.** Each point represents the p-value for the optimal subset for a given SNP. Blue SNPs are genome-wide significant SNPs in loci that had previously been identified in one of the original GWASs. Dark orange SNPs represent genome-wide significant SNPs in novel loci.

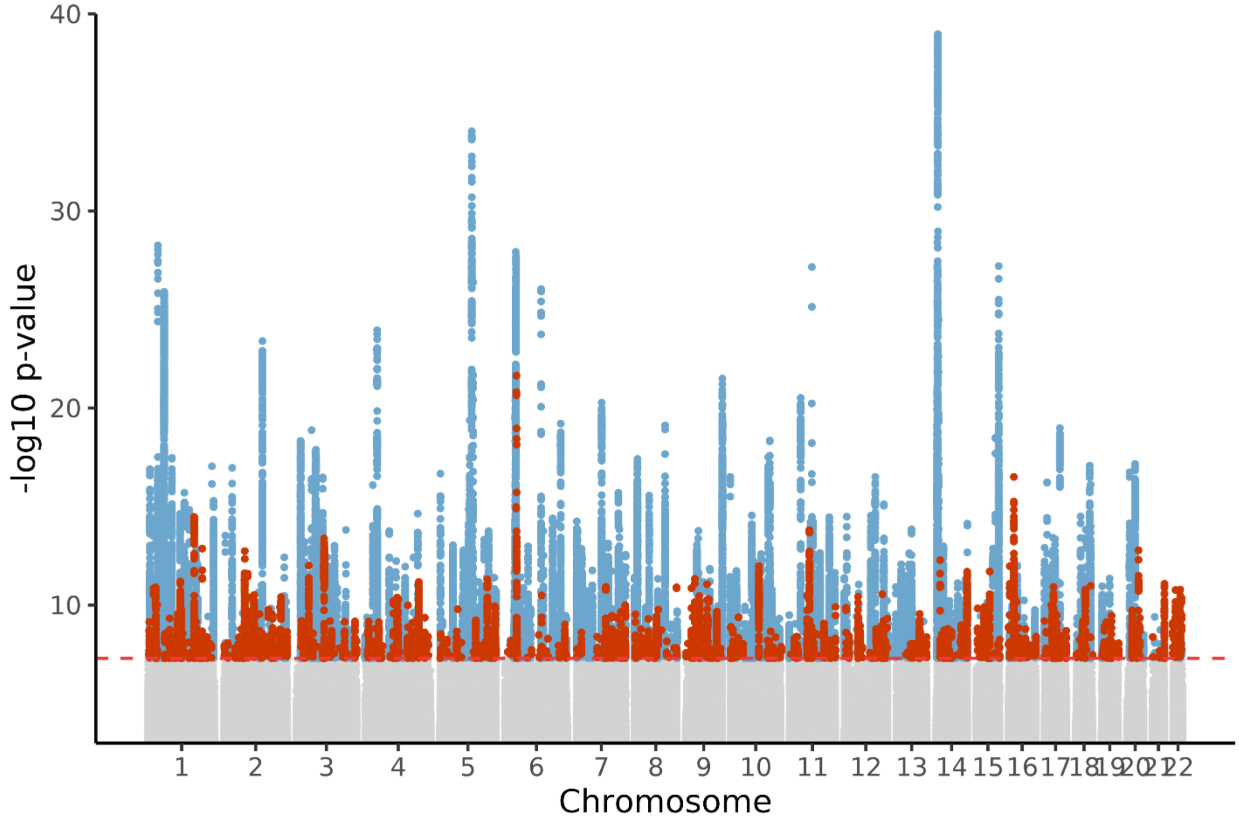

**Supplementary Figure 9. A Manhattan plot representing the CPMA results for cross-trait associations across psychiatric traits**. Each point represents the p-value for the optimal subset for a given SNP. Blue SNPs are genome-wide significant SNPs in loci that had previously been identified in one of the original GWASs. Dark orange SNPs represent genome-wide significant SNPs in novel loci.

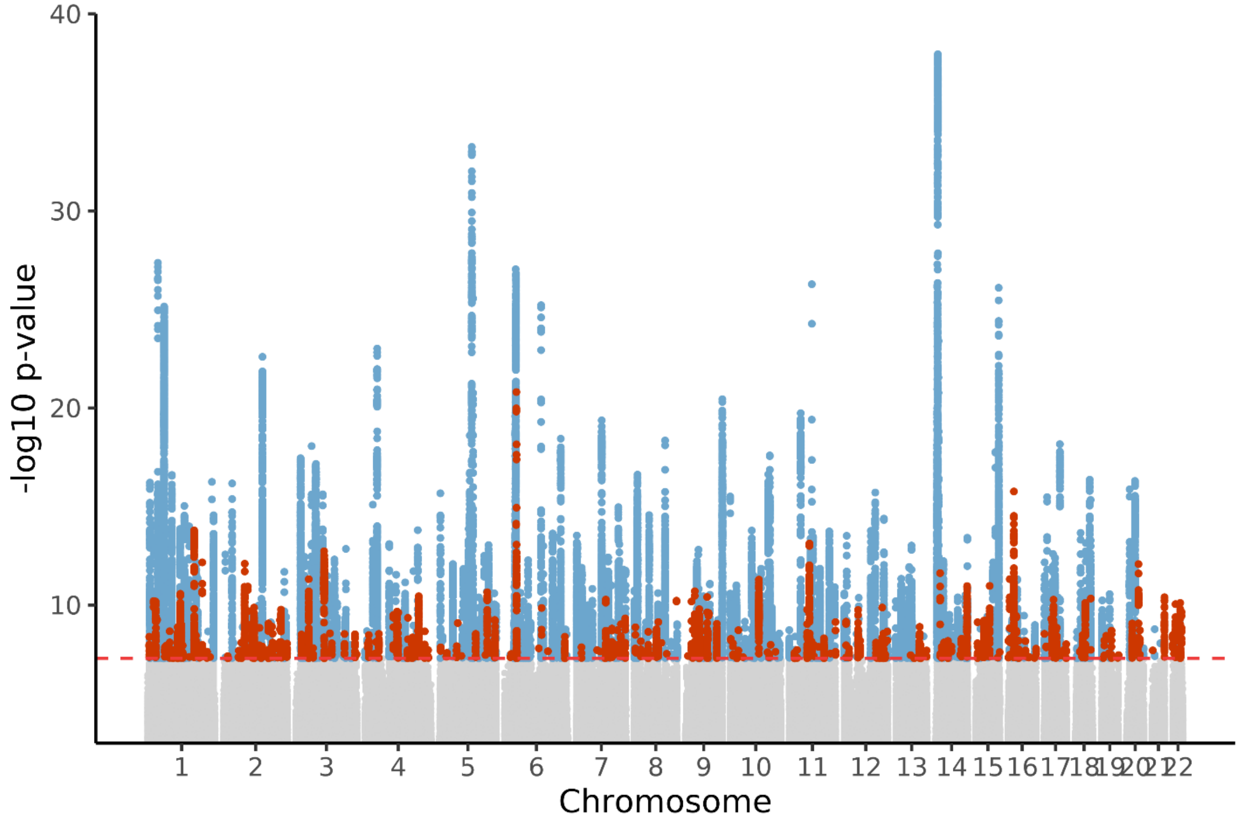

**Supplementary Figure 10. A Manhattan plot representing the ASSET results for cross-trait associations across psychiatric traits**. Each point represents the p-value for the optimal subset for a given SNP. Blue SNPs are genome-wide significant SNPs in loci that had previously been identified in one of the original GWASs. Dark orange SNPs represent genome-wide significant SNPs in novel loci.

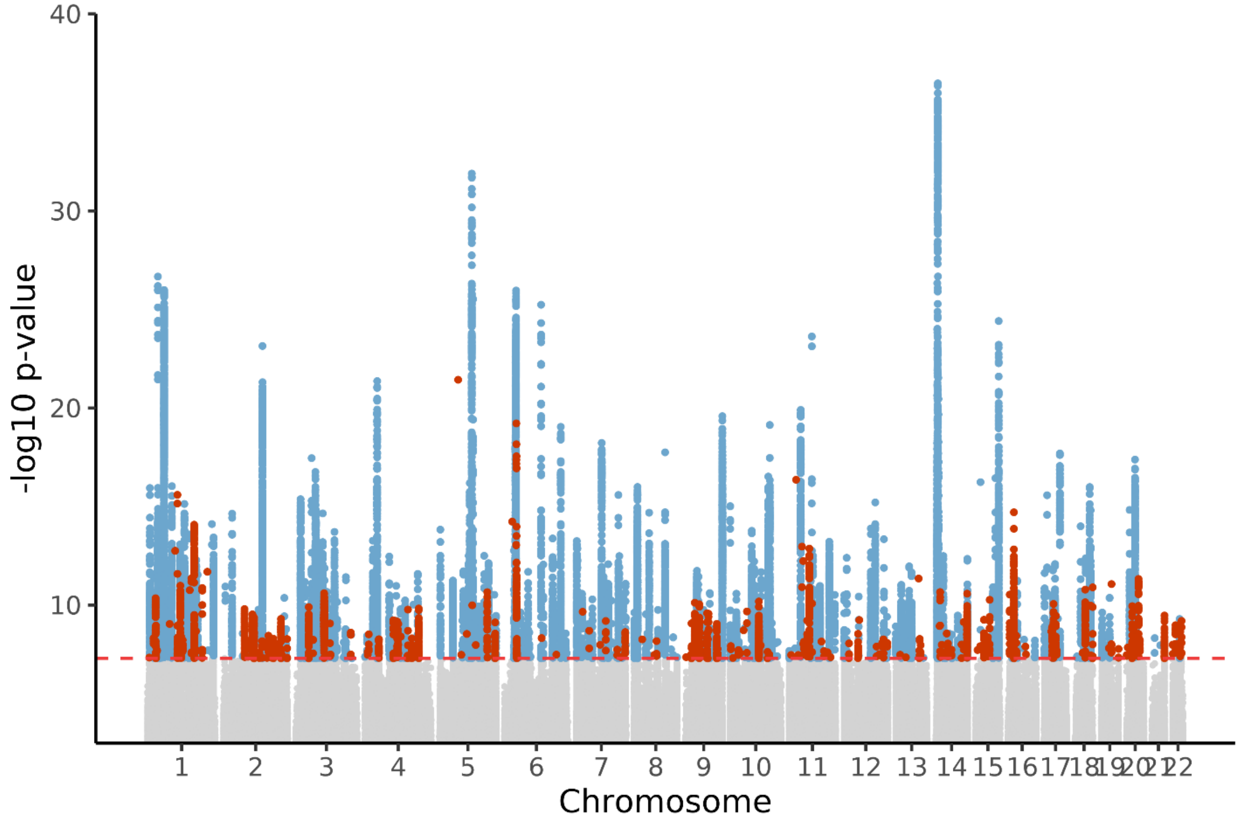

**Supplementary Figure 11. Violin plots of the basewise length of genome-wide significant loci for each of the cross-trait association methods**.

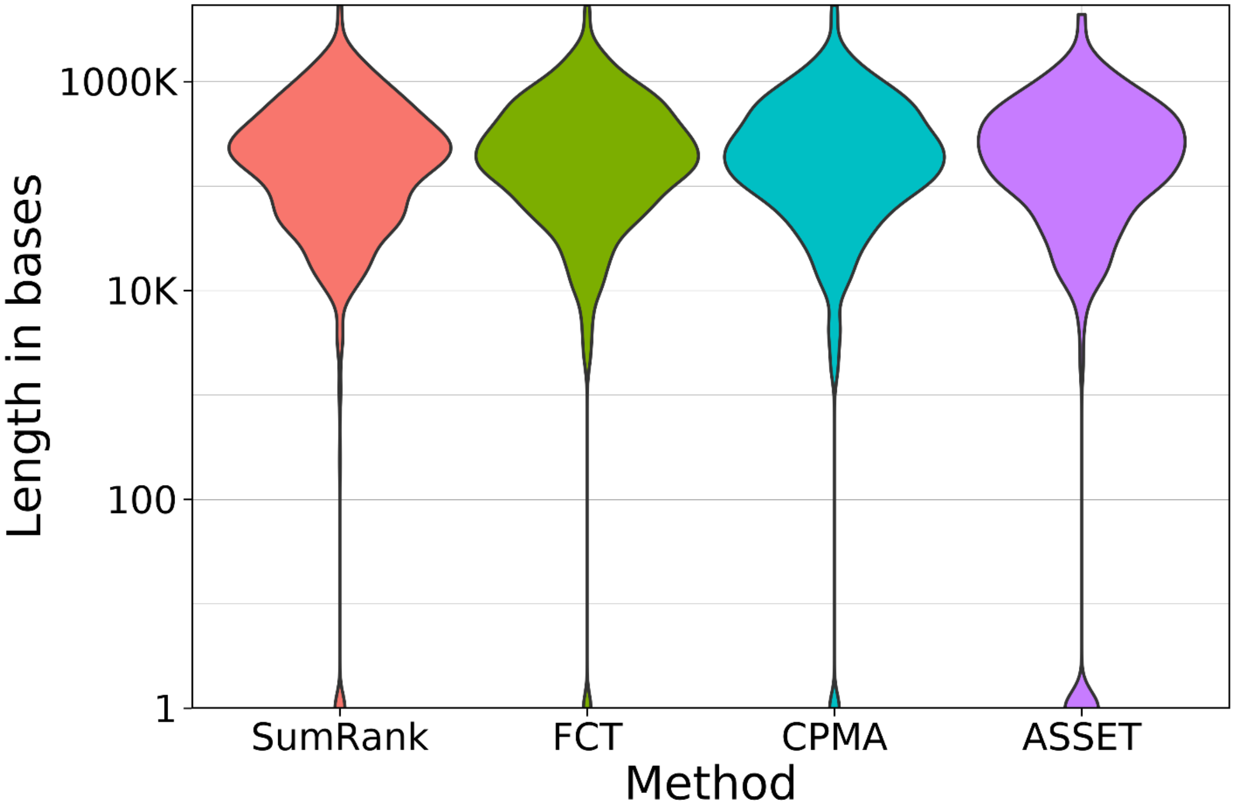

**Supplementary Figure 12. The functional categories for SNPs identified by SumRank, FCT, CPMA, and ASSET.**

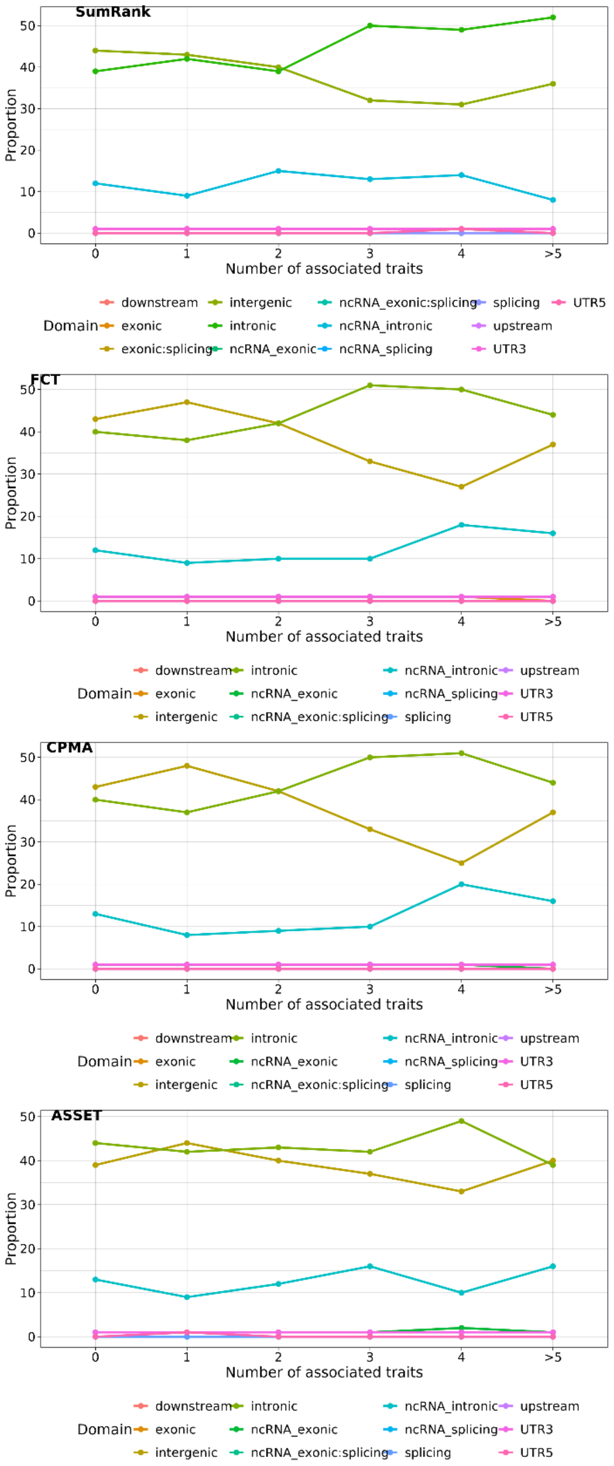

**Supplementary Figure 13. Tissue-specificity analysis results for the cross-trait association methods**. The results are shown for SumRank (top-left), FCT (top-right), CPMA (bottom-left), and ASSET (bottom-right).

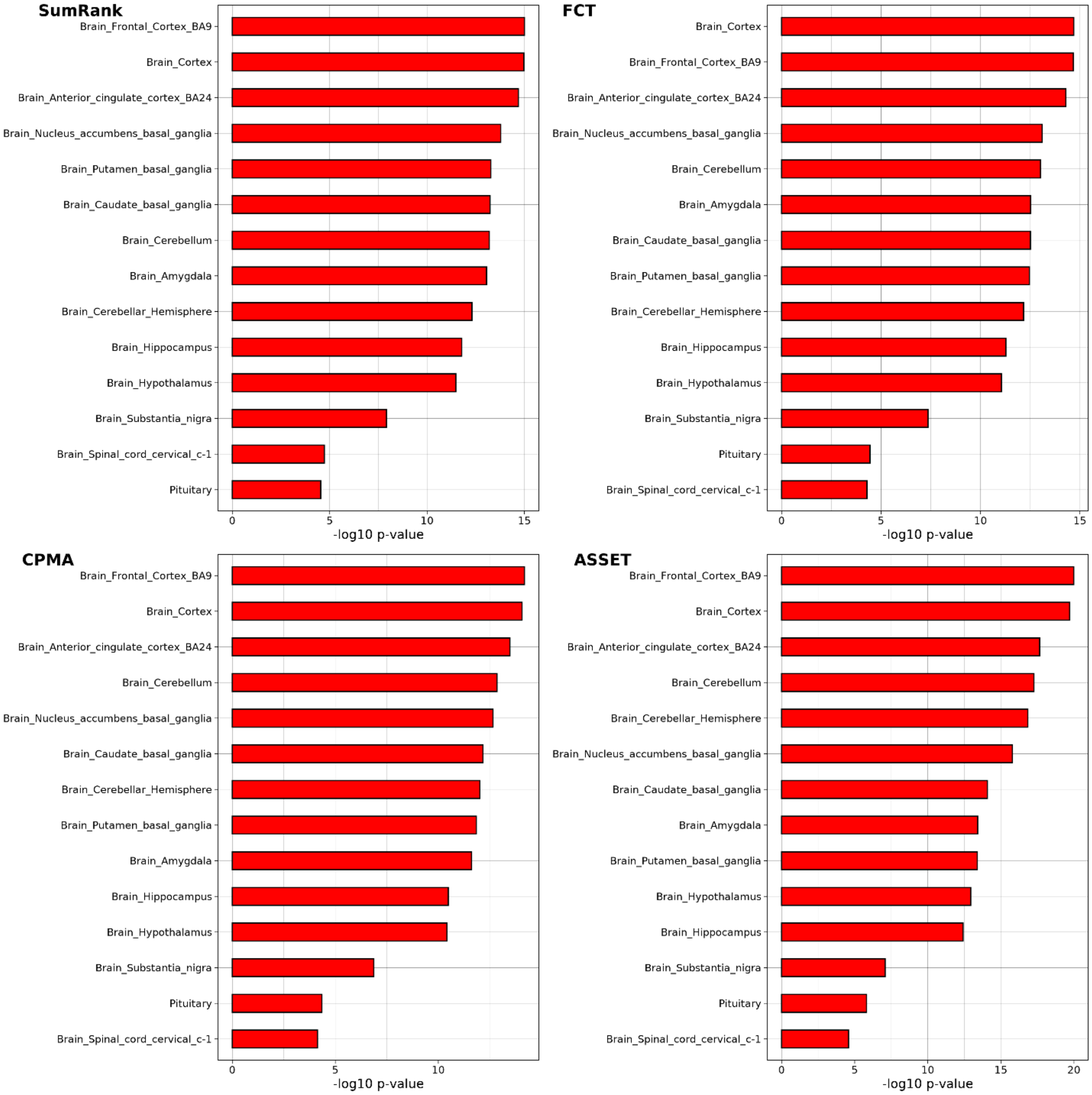
